# Clinical sensitivity and specificity of a high-throughput microfluidic nano-immunoassay combined with capillary blood microsampling for the identification of anti-SARS-CoV-2 Spike IgG serostatus

**DOI:** 10.1101/2022.06.09.22276142

**Authors:** Grégoire Michielin, Fatemeh Arefi, Olha Puhach, Mathilde Bellon, Pascale Sattonnet-Roche, Arnaud G. L’Huillier, Isabella Eckerle, Benjamin Meyer, Sebastian J. Maerkl

## Abstract

**Background:** We evaluate the diagnostic performance of dried blood microsampling combined with a high-throughput microfluidic nano-immunoassay (NIA) for the identification of anti-SARS-CoV-2 Spike IgG seropositivity.

**Methods:** We conducted a serological study among 192 individuals with documented prior SARS-CoV-2 infection and 44 SARS-CoV-2 negative individuals. Participants with prior SARS-CoV-2 infection had a long interval of 11 months since their qRT-PCR positive test. Serum was obtained after venipuncture and tested with an automated electrochemiluminescence anti-SARS-CoV-2 S total Ig reference assay, a commercial ELISA anti-S1 IgG assay, and the index test NIA. 109 participants from the positive cohort and 44 participants from the negative cohort also participated in capillary blood collection using three microsampling devices: Mitra, repurposed glucose test strips, and HemaXis. Samples were dried, shipped by regular mail, extracted, and measured with NIA.

**Findings:** Using serum samples, we achieve a clinical sensitivity of 98·33% and specificity of 97·62% on NIA, affirming the high performance of NIA in participants 11 months post infection. Combining microsampling with NIA, we obtain a clinical sensitivity of 95·05% using Mitra, 61·11% using glucose test strips, 83·16% using HemaXis, and 91·49% for HemaXis after automated extraction, without any drop in specificity.

**Interpretation:** High sensitivity and specificity was demonstrated when testing micro-volume capillary dried blood samples using NIA, which is expected to facilitate its use in large-scale studies using home-based sampling or samples collected in the field.

**Funding:** Swiss National Science Foundation NRP 78 Covid-19 grant 198412 and Private Foundation of the Geneva University Hospital.

**Research in context:** *Evidence before this study:* Serological surveillance is of importance to better understand the evolution and spread of SARS-CoV-2 and adapt public health measures. We identified multiple studies conducting such serological surveys using decentralized collection of capillary blood, facilitating the logistics and reducing burden on participants and healthcare facilities. To perform the detection of anti-SARS-CoV-2 antibodies with a high-throughput and at low-cost, a microfluidic nano-immunoassay (NIA) was developed which requires ultra-low sample volumes and minimizes reagent consumption.

*Added value of this study:* In this study we showed the possibility of combining capillary microsampling with NIA. We validated the use of NIA in serum samples obtained 11 months after infection and show the good clinical performance of the assay in samples with waning antibody titers. Using three different microsampling device, namely Mitra, repurposed glucose test strips, and HemaXis, we implemented a protocol using dried blood sample collection, shipping, extraction, and testing on the microfluidic assay. The sensitivity and specificity were measured and are presented when using the different microsampling devices.

*Implications of all the available evidence:* We show that the performance of NIA is good when using serum samples, but also in combination with microsampling. Facilitated logistics and increased convenience of microsampling, together with high-throughput and low-cost testing on a microfluidic assay should facilitate the conduction of serological surveys.

## Introduction

After its appearance in 2019, severe acute respiratory syndrome coronavirus 2 (SARS-CoV-2) rapidly caused a global pandemic of coronavirus disease of 2019 (COVID-19), totaling over 500 million declared cases as of spring 2022[1]. Serosurveillance was quickly recognized as an important tool to better understand the evolution of the pandemic and inform public health decisions[2]. The first published results for SARS-CoV-2 seroprevalence studies were from Stringhini *et al*. describing the seroprevalence after the first wave of the pandemic in the population of Geneva[3]. Also, a coordinated nationwide serosurveillance program called Corona Immunitas helps gather critical data on the SARS-CoV-2 epidemiological situation in Switzerland[4]. Similar serological surveillance programs were conducted in France[5], Netherlands[6], India[7], and the United States of America[8], among others[9].

In order to facilitate such serological surveys, and in particular when proposing longitudinal or repeat testing, it is critical to reduce the burden on study participants as much as possible by simplifying blood sampling and avoiding unnecessary visits to testing centers, while less invasive sampling is especially important when including children into such surveys. However, many serological surveys still rely on a visit to a healthcare center or testing facility where venous blood draws are obtained. Visits to central care facilities could present the participants and staff with a risk of infection. Moreover, some populations are difficult to include during an epidemic or pandemic when such serosurveys are of utmost importance, due to their limited access to proper facilities or due to possible vulnerabilities, such as persons living in elderly care facilities. When large-scale serosurveillance programs are conducted, costs associated with sampling, sample preparation, samples shipping, and analysis also become important parameters.

Capillary blood microsampling offers the possibility to decentralize blood collection by untrained individuals, and there is evidence of successful use of capillary blood samples in SARS-CoV-2 serological surveys. Indeed, self-collected capillary blood samples can be sent directly in a liquid format using microtubes[10] or using specific collection devices such as Tasso-SST[11, 12], however the collected volumes are generally higher compared to microsampling at around 250-500 *µ*L. Also, facilitated logistics by shipping at room temperature with long-term stability and reduced biohazard can be achieved after drying of blood samples, while still allowing appropriate detection of anti-SARS-CoV-2 antibodies. For example, Beyerl *et al*. used at-home blood spot collection on standard filter paper cards in a large-scale sero-survey[13]. They obtained high sensitivity and specificity when extracting three sub punches of the dried blood spots and assaying the eluate in micro sample cups using the Elecsys anti-SARS-CoV-2 N total Ig assay on the Roche system.

Microsampling devices obtaining a defined blood volume have also been used, with the potential advantage of removing the hematocrit bias associated with the processing of samples directly spotted on filter paper[14]. For instance, direct collection of capillary blood microsamples on Neoteryx Mitra devices has been shown to provide comparable results to serum samples using the Roche Elecsys assay[15] or in combination with ELISAs[16, 8]. Similarly, studies showed the potential of using other volumetric microsampling systems by using minivette and subsequent transfer on filter paper[17], or using devices which, like Mitra, also allow direct microsample collection of defined volume such as Hemapen[18], Capitainer[19] or HemaXis[20].

While the performance of dried blood sample testing on existing assays is good, sample dilution upon extraction can lead to problems of lowered sensitivity[21]. Also, the small volume of the eluate can be a limitation for reanalysis, for subsequent testing of other parameters, or for biobanking applications.

To address these limitations, we previously demonstrated the possibility to detect the presence of SARS-CoV-2 anti-spike IgG antibodies in ultralow-volume blood samples using a microfluidic nano-immunoassay (NIA) validated on 289 serum samples[22]. The platform allows for high-throughput testing of up to 1024 samples in parallel on a single device, with minimal sample or reagent consumption, leading to cost-effective serological testing, and excellent assay performance with 100% specificity and 98% sensitivity. As a proof-of-concept, we also showed the compatibility of NIA with low-volume dried blood samples by transferring EDTA whole blood on microsampling devices, followed by drying, extraction, and testing with NIA.

In this study, we aimed to quantify the performance of NIA (index test) in analyzing capillary blood samples collected on three different microsampling devices, namely Neoteryx Mitra, repurposed glucose test strips, and DBS Systems HemaXis. We set out to determine whether NIA can be used to measure SARS-CoV-2 anti-Spike IgG antibody levels in dried capillary blood microsamples and to assess and compare the so obtained sensitivity and specificity with serum samples from the same individuals analyzed by Roche Elecsys anti-SARS-CoV-2 S total Ig, Euroimmun ELISA anti-S1 IgG, and NIA.

We show the ability of NIA to classify subjects with prior SARS-CoV-2 infection 11 months post infection documented by qRT-PCR (SARS-CoV-2 positive) and subjects without known prior infection (SARS-CoV-2 negative) using serum obtained by venipuncture. Next, we present the performance of the assay when used in combination with microsampling using Mitra, repurposed glucose test strips, or HemaXis, and also present automated dried blood spot (DBS-A) extraction of HemaXis. Using serum samples, NIA shows excellent performance with clinical sensitivity of 98·33% and specificity of 97·62% in samples taken 11 months post infection. Combined with microsampling, there is no change in specificity on NIA and the sensitivity remains high at 95·05% when using Mitra device, 61·11% using glucose test strips, 83·16% with HemaXis, and 91·49% for HemaXis after DBS-A extraction. We compare the results with matching serum samples measured on the Roche Elecsys anti-SARS-CoV-2 S total Ig as reference assay and show a 98·20% agreement when measuring serum samples on the microfluidic NIA index test, as well as a 95·80% agreement when combined with microsampling on Mitra, 72·55% with glucose test strips, 87·22% with HemaXis, and 93·75% after DBS-A extraction of HemaXis. Together, these results indicate a good performance of NIA when using dried capillary blood samples and validate its utility in performing serological surveys, allowing decentralized sample collection and centralized analysis with high-throughput, low sample volume requirements, and low reagent consumption.

## Materials and methods

### Participants and study design

We did not perform a sample size calculation. We aimed at obtaining over 100 participants in the SARS-CoV-2 positive group and over 50 participants in the SARS-CoV-2 negative group based on participants availability. This would allow a lower limit of the 95% confidence interval above 85% in the sensitivity and specificity evaluation[23], assuming equivalent sensitivity and specificity as our previous results based on serum analysis[22]. Adult participants with a documented PCR-positive SARS-CoV-2 infection were eligible in the positive group, and participants without knowledge of previous SARS-CoV-2 infection were eligible in the negative group. The patients in the positive group were already identified and part of another study[24], and had a documented previous infection 6-12 months before the start of this study. Between February and March 2021, 200 participants were invited to participate in a follow-up serology using venous blood as part of ongoing studies, and were solicited for their participation in this additional investigation using capillary blood microsampling. Participants in the negative group, which were recruited specifically for this study, consisted of a convenience sample of non-clinical employees of the University of Geneva or Geneva University Hospital. The corresponding study diagram in Figure 1 was created with the Lucidchart web application. This study was approved by the local ethics committee (CCER project numbers 2020-00516 and 2020-02323) and registered (NCT04329546) prior to initiation.

**Figure 1:**
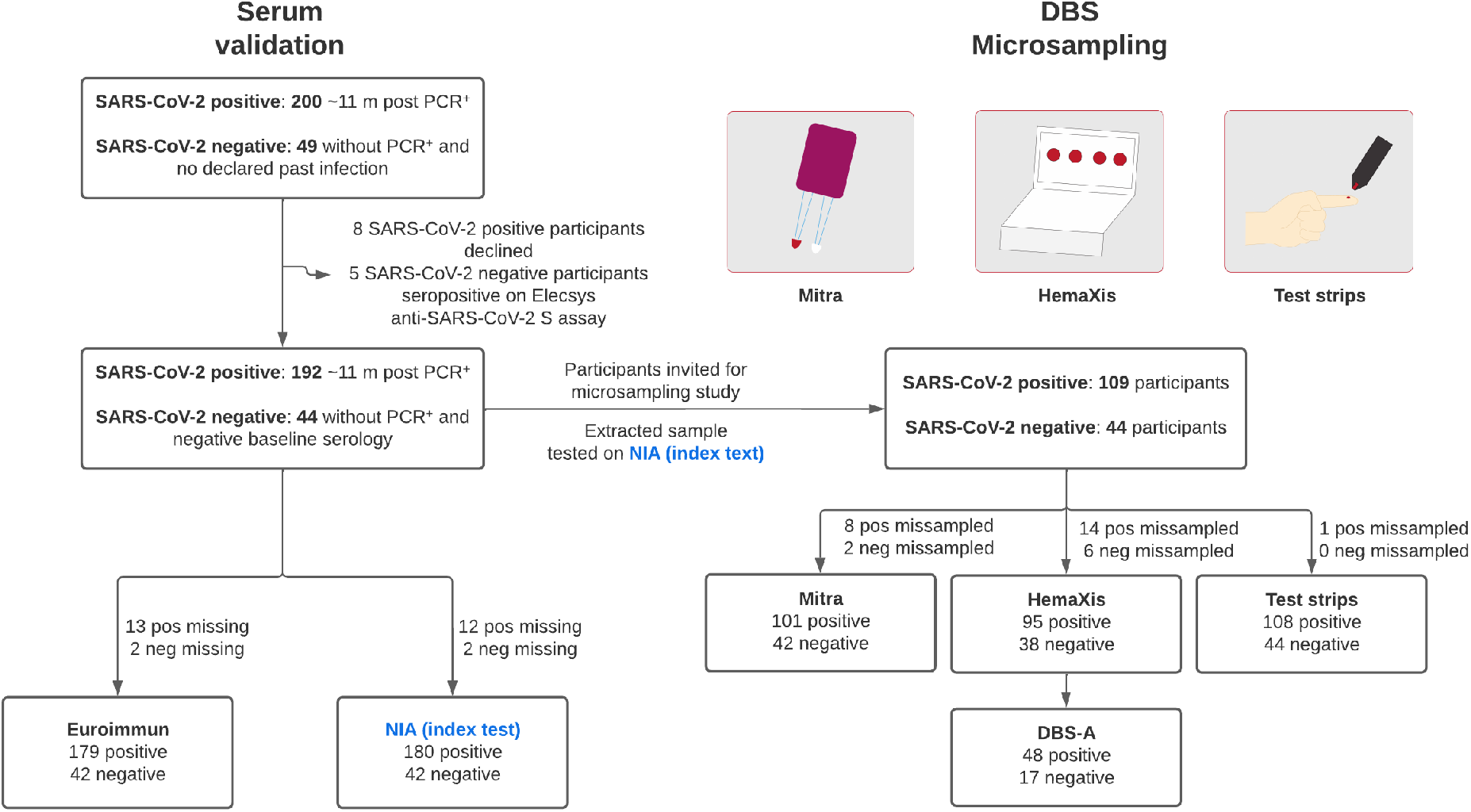
**Study participants. Serum validation)** participants were included in this study and were invited for a venipuncture. Serum obtained was used for testing on the reference standard Roche Elecsys anti-SARS-CoV-2 S total Ig assay, Euroimmun anti-S1 IgG assay, and on the microfluidic nano-immunoassay (NIA) index test. **DBS microsampling)** On the same day, the participants were invited to perform a capillary blood collection using three different microsampling devices: Mitra, repurposed glucose test strips, and HemaXis. The samples were shipped by regular mail in a dried state and extracted upon reception. For filter cards collected with the HemaXis device, a subset of samples was extracted by flow-though desorption using an automated instrument (DBS-A, Gerstel AG).

### Sample collection and preparation

#### Serum samples

Serum samples obtained from venipuncture were available for participants as part of the investigations of the persistence of anti-SARS-CoV-2 antibodies[24]. Aliquots were retrieved from the existing blood bank, heat inactivated at 56^°^C and Triton X-100 was added to each sample to a concentration of 1%. The inactivated samples were shipped refrigerated by post from Geneva to Lausanne for analysis.

#### Capillary blood microsampling

Each capillary microsampling kit contained: general material for capillary blood sampling (3 lancets (BD Microtainer High Flow - 21 G 2·0 mm), pre injection cleansing swab (PDI,70% isopropyl alcohol), gauze, plasters, return sample bag with dessicant, return box), 1 Mitra sampling device in cartridge format with 2 × 10 *µ*L sample collectors in a resealable sample bag containing dessicant (Neoteryx), 1 Hemaxis device with 4 × 10 *µ*L sampling channels (DBS System SA), and 2 × 0·6 *µ*L glucose test strip (Medisana MediTouch 2) placed in 2 mL tubes with pierced cap to allow sample drying. With the exception of one participant who collected the samples at home, all samples were collected at the Geneva University Hospital with the supervision and help of research staff. After de-identification, samples were sent by post every week in a package containing samples from the previous week.

#### Sample processing

To minimize the risk of exposure to biohazardous agents, the samples were extracted after more than 4 days in a dried state. The samples were extracted in a buffer containing surfactants, namely phosphate buffered salined (PBS) containing 1% bovine serum albumin (BSA) and 0·5%Tween-20 (PBS-BT)[16]. The Mitra tips were separated from the stem and Hemaxis spots on paper cards were punched out with an 8 mm puncher, and were each placed in a well of a 96-well plate and 200 *µ*L PBS-BT was added to each well (10 *µ*L sample = 20x dilution factor). The plates were incubated at 37^°^C for 3h on a ThermoMixer C with 300 rpm agitation and a ThermoTop lid to prevent condensation (Eppendorf). For glucose test strips, the 0·6 *µ*L samples were extracted in 30 *µ*L PBS-BT (50x diluton factor) in order for the test strip to be sufficiently immersed in the extraction buffer at the bottom of the 2 mL tube, and were incubated at 37^°^C for 3h without agitation. All extracted samples were stored at −20^°^C until analysis.

#### Automated DBS extraction

For the automated extraction of DBS on filter paper cards, we used a dried blood spot autosampler[25] (Gerstel SA). A 6mm clamp was mounted to extract samples by flow-through desorption using PBS-BT, and 2 × 150 *µ*L were flowed at a speed of 700 *µ*L/min through the spot allowing collection of 2 fractions. To improve spot desorption, the extraction buffer was briefly heated to 80^°^C before flowing through the spot. The first fraction was discarded as it corresponded to air void volume or washing buffer contained in the tubing after the clamp. The second fraction was collected in a 96-well round-bottom microtiter plate (Nunc) corresponding to a volume of around 80 *µ*L with the rest of the 150 *µ*L remaining in the tubing and eliminated in subsequent washing steps. Between samples, a washing of all the tubing and lines was performed with 1000 *µ*L of extraction buffer at a speed of 4000 *µ*L/min.

### Microfluidic nano-immunoassay

#### Microfluidic chip fabrication

For the microfluidic chip fabrication, we used the same protocol as described previously without modification[22].

#### Microarray spotting

The extracted samples were thawed on ice and 25 *µ*L were transferred to a low-volume 384-well plate (Arrayit, MMP384). Each sample was deposited into 4 spots using a randomized spotting pattern, and two stamps per spot with an inking time of 50 ms and printing time of 1 ms. Other microarray spotting parameters were kept as described previously[22].

#### Immunoassay reagents

We used the same reagents as described in our previous study[22]. BSA-biotin (Thermo Fisher, 29130) and neutrAvidin (Thermo Fisher, 3100) were used for surface modification of the microfluidic device. We used biotinylated mouse anti-His antibodies (Qiagen Cat# 34440, RRID:AB 2714179) followed by full length His-tagged SARS-CoV-2 Spike produced at the EPFL protein facility. The prefusion ectodomain of SARS-CoV-2 spike glycoprotein (the construct was a generous gift from Prof. Jason McLellan, University of Texas, Austin(4)) was transiently transfected into suspension-adapted HEK293 cells (Thermo Fisher) with PEI MAX (Transfection grade linear polyethylenimine hydrochloride, Polysciences) in Excell293 medium. Incubation with agitation was performed at 37^°^C and 4·5% CO2 for 5 days. The clarified supernatant was loaded onto Fastback Ni2+ Advance resin column (Protein Ark) eluted with 500 mM imidazole, pH 7·5 in PBS. For detecting human IgG, we used PE labeled goat anti-Human-IgG (Abcam Cat# ab131612, RRID:AB 11156857).

#### Running on-chip immunoassays

We used the same procedure described in a previous study when performing the microfluidic nano-immunoassay[22]. Briefly, the microfluidic assay allows the succesive flowing of immunoassay reagents and resolubilization of spotted sample which are individually assayed in a unit cell. The unit cells were imaged using an automated microscope stage. In addition to a fluorescent image, a brightfield image was acquired for each unit cell at the same coordinates for use in the image analysis.

#### Image analysis

We used FIJI and a custom macro to process the image stacks for the 1024 unit cells. Using the brightfield image, a circular region corresponding to the MITOMI valve area was detected using a Hough Circle Transform plugin (UCB Vision Sciences library). An ROI defined as the button area was obtained after 30 steps of erosion of the MITOMI valve region. A second ROI defined as the background area was obtained after 30 dilation steps of the MITOMI valve region, and substracted by the button area ROI. The final signal for each unit cell was calculated as the median of the button area substracted by the median of the background area. For each sample, the mean and standard deviation of 4 technical repeats was calculated.

#### Selection of reference assay

The Roche anti-SARS-CoV-2 S RBD total Ig was selected because of its reported high sensitivity and specificity by the manufacturer, and it was also previously shown to achieve high sensitivity in subjects with long interval since time of infection[26]. The Euroimmun anti-SARS-CoV-2 S1 IgG ELISA was also selected for comparison as it was the first CE-marked serological assay targeting part of the SARS-CoV-2 spike antigen[27] and was extensively validated in the Geneva Center for Emerging Viral Diseases[28].

#### Roche Elecsys anti-SARS-CoV-2 S RBD total Ig

SARS-CoV-2 specific antibodies were determined using the quantitative Elecsys S RBD total Ig assay on a cobas e801 analyser (Roche Diagnostics, Rotkreuz, Switzerland) in the clinical laboratory of the University of Geneva Hospital. Results are reported as concentrations (U/mL) and positivity was determined by using the manufacturer’s cuf-off >0·8 U/mL.

#### Euroimmun anti-S1 IgG

Euroimmun anti-SARS-CoV-2 S1 IgG ELISA (Euroimmun AG, Lübeck, Germany # EI 2606-9601 G) was performed manually according to the manufacturer’s instructions. The reactivity of each sample was measured at an optical density of 450nm (OD450) and then divided by the OD450 of the calibrator provided with each ELISA kit to minimize inter-assay variation. The quantitative results obtained were then expressed in arbitrary units and interpreted as follows: OD ratio: <0·8 = negative; ≥0·8 and <1·1 = borderline; ≥1·1 = positive.

#### Statistics

Statistical analyses were performed using GraphPad Prism Version 9.3.1. Every sample was tested in four technical replicates and the mean was used in ROC analysis. The threshold for positivity was selected as the maximum likelihood ratio calculated and the corresponding sensitivity and specificity with 95% confidence interval were obtained. The receiver operating characteristic curve and area under the curve with 95% confidence interval was also calculated. To estimate the concordance with the Roche assay, Kappa coefficient calculation was done using the online tool available at https://www.graphpad.com/quickcalcs/kappa1/. For the reproducibility between two microfluidic experiments, the mean and standard deviation of four technical replicates on each experiment was used, and a simple linear regression was performed using GraphPad Prism. For the comparison between serum and the different microsampling methods, a log transform of the mean was calculated and a simple linear regression was performed using GraphPad Prism.

#### Ethics statement

Ethical approval for this study was given by the Ethics Committee Geneva (Commission Cantonale d’Ethique de la Recherche sur l’être humain (CCER) project numbers 2020-00516 and 2020-02323) and registered (NCT04329546) prior to initiation.

#### Role of the funding sources

The funding sources did not play any role in the study design, in the collection, analysis and interpretation of the data, in the writing of the report, or in the decision to submit the paper for publication.

## Results

### Participants

The description of the study is presented in Figure 1. We invited 200 participants enrolled in an ongoing longitudinal serological study using venous blood with documented SARS-CoV-2 infection by positive qRT-PCR[24], and 192 (96%) participated in this study consisting in a follow-up visit 11 months post infection (SARS-CoV-2 positive group). In addition to a venipuncture, 109 subjects (56·77%) agreed to take part in the microsampling study. The subjects in the SARS-CoV-2 negative group consisted of 49 individuals without known past infection and were recruited specifically for this study. 5 participants in the SARS-CoV-2 negative group who tested positive on Elecsys anti-SARS-CoV-2 S assay were excluded from the study. All 44 participants in the SARS-CoV-2 negative group had venipuncture and microsampling performed. Participants in the cohort had a median age of 41·21 (interquartile range: 30·98-50·98) and 174/249 (69·86%) were women.

### Blood sampling

For capillary blood microsampling, participants were provided with two commercially available microsampling devices: Neoteryx Mitra and DBS System HemaXis. We also tested repurposed glucose test strips as a low-cost, ultra-low volume alternative (Fig. 1). Microsampling was performed on the same day as the venipuncture, and 153 individuals participated in micro-volume capillary blood sampling (62·70% of the total). Each participant received a kit containing all microsampling devices and three lancets to ensure the possibility of repeating a fingerprick if needed between the different collections. Participants received the suggestion to first collect capillary blood samples on the Mitra device, followed by the glucose test strips and then the HemaXis device, and we note that this order could influence sample collection success rate.

Based on visual inspection of the collection tip for presence or absence of blood, as well as appropriate sampling according to the instructions for use provided by the manufacturer, the Mitra device allowed for 143 (93·46%) successful collections on at least one of the two collection tips (2 samplers per Mitra cartridge). Among the 10 failed sampling events, 8 were excluded due to insufficient sampling, and 2 were due to oversampling manifested by blood present on the stem of the Mitra sampler. The glucose test strips allowed collection of blood in 152 (99·35%) cases, with one participant who did not collect any visible blood. We note that it was not possible to visually assess whether the 0·6 *µ*L test strip channel was under- or over-sampled. Therefore any glucose test strip with visible blood collection was considered as a successful collection. Sample collection with the Hemaxis device was successful in 133 (86·93%) of cases, with participants having at least one correctly sampled blood spot. 22 samples had no dried blood spot or had spots of small size due to undersampling, while oversampling was not observed. As each HemaXis device contains 4 available collection channels, 65 participant (42·48%) could collect an additional third or fourth spot which were used for automated extraction using flow-through desorption (DBS-A instrument, Gerstel AG). These results show that microsampling of capillary blood results in a high sampling success rate, even when collection on multiple different devices was required.

### Sample validation in serum

We used serum samples obtained by venipuncture for measurements on the reference Roche Elecsys anti-SARS-CoV-2 S total Ig assay, on Euroimmun ELISA anti-S1 IgG assay, and on NIA for initial validation. We tested a total of 192 samples from subjects with prior SARS-CoV-2 infection (SARS-CoV-2 positive) and 44 samples from subjects without prior infection (SARS-CoV-2 negative) with the Elecsys anti-SARS-CoV-2 S total Ig reference assay, 179 SARS-CoV-2 positive and 42 SARS-CoV-2 negative with the ELISA anti-S1 IgG assay, and 180 SARS-CoV-2 positive and 42 SARS-CoV-2 negative on the NIA assay(Fig. 1). The difference in the number of samples tested is explained by serum samples being initially tested on the reference Elecsys assay, while samples were retrieved at a later timepoint only when sufficient volume was available for the measurement on ELISA and NIA assays.

For the Elecsys anti-SARS-CoV-2 S total Ig reference assay of serum samples (Fig. 2A), all 192 participants with available sample of the SARS-CoV-2 positive cohort showed a signal above the positivity threshold (≥0·8 U/mL). 8 samples had signal >5000 U/mL and the 184 remaining samples signal had a geometric mean of 140·4 U/mL (95% CI: 116·7-169·0) with an interquartile range of 68·03-343·8 U/mL.

**Figure 2:**
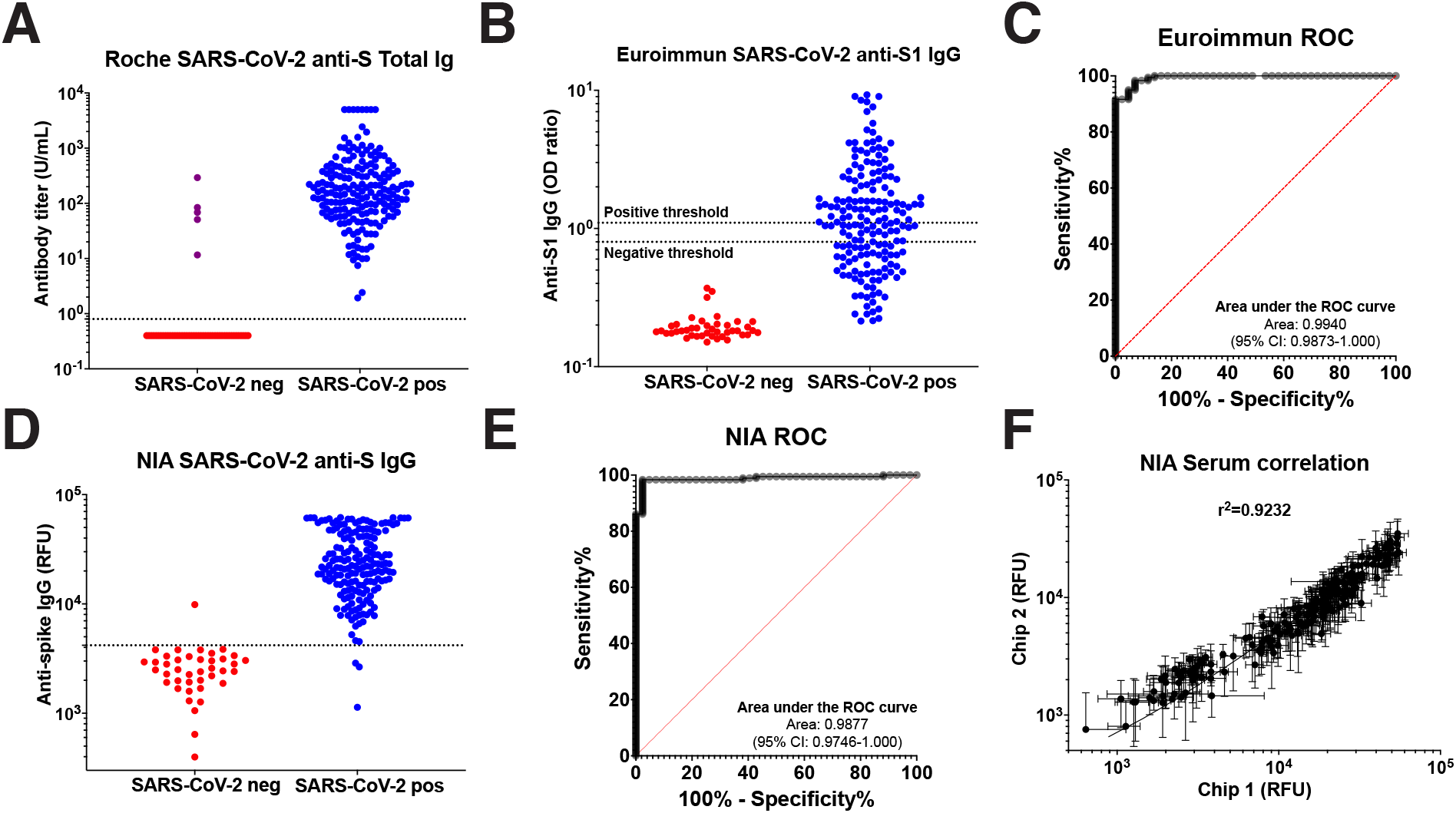
Sample validation with Roche Elecsys, Euroimmun ELISA, and NIA. **A)** Roche Elecsys anti-SARS-CoV-2 S total Ig assay. 5 SARS-CoV-2 negative samples (purple color) with signal above the positivity threshold (>0·8) were excluded from the study. **B)** Euroimmun ELISA anti-S1 IgG assay. The thresholds for positivity (>1·1) and negativity (<0·8) classification as provided by the manufacturer are indicated. **C)** ROC curve with calculated AUC for Euroimmun ELISA. **D)** NIA measuring inactivated serum samples at 1:8 dilution. **E)** ROC curve with calculated AUC for NIA. **F)** Correlation between two different NIA chips (N=198). Samples with saturating signals (>55’000 RFU) were removed from the plot and correlation calculation.

For the Euroimmun ELISA anti-S1 IgG assay, we observe a relatively poor discrimination between SARS-CoV-2 positive and SARS-CoV-2 negative samples (Fig. 2B). Based on the positivity threshold provided by the manufacturer (≥1·1), the ROC analysis returns a clinical sensitivity of 51·40% (95% CI: 44·12-58·61) and specificity of 100% (95% CI: 91·62%-100·0), resulting in a low concordance with the Roche reference assay of 60·63% observation agreement and a Kappa coefficient of 0·287 (95% CI: 0·205-0·369). Considering an approximate prevalence of 25% at that stage in the pandemic[29], this would correspond to a positive predictive value (PPV) of 100%, a negative predictive value (NPV) of 86·06% and a diagnostic accuracy of 87·85%. Finally, the receiver operating characteristic curve (AUROC) is 0·9940 (95% CI: 0·9873-1·000) (Fig. 2C).

The microfluidic NIA test showed excellent discrimination between SARS-CoV-2 positive and SARS-CoV-2 negative samples when analyzing heat-inactivated serum at a 1:8 dilution (Fig. 2D). The AUROC is 0·9877 (95% CI: 0·9746-1·000) (Fig. 2E) and the positivity threshold determined by the maximum likelihood (>4182 RFU) gives a clinical sensitivity of 98·33% (95% CI: 95·22-99·55) and specificity of 97·62% (95% CI: 87·68-99·80). This result is similar to the clinical sensitivity observed in our previous study[22], and demonstrates the robustness of NIA in detecting the presence of anti-SARS-CoV-2 antibodies not only in subjects with recent infection, but also following a long interval of 11 months between infection and serological testing. The percentage agreement with the Roche assay was 98·20% with a Kappa coefficient of 0·942 (95% CI: 0·886-0·998). With a prevalence of 25%, this would correspond to a PPV of 93·23%, a NPV of 99·43% and a diagnostic accuracy of 97·79%. We note that only three samples in the SARS-CoV-2 positive cohort have NIA signals below the threshold, while for the SARS-CoV-2 negative cohort a single sample has a positive signal on NIA leading to the observed specificity below 100%. Finally, we evaluated the reproducibility between measurements performed on two different days on NIA (N=198) and observed a correlation with an R^2^ = 0·9232 (Fig. 2F).

### Mitra microsampling

Combining Mitra capillary blood collection with NIA resulted in very good discrimination between SARS-CoV-2 positive and SARS-CoV-2 negative samples (Fig. 3A). The AUROC is 0·9892 (95% CI: 0·9754-1·000) (Fig. 3B) and we determined a threshold for positivity using the maximum likelihood (>2089 RFU), which returns a clinical sensitivity of 95·05% (95% CI: 88·93-97·87) and specificity of 97·62% (95% CI: 87·68-99·88). The Mitra device gave the highest clinical sensitivity among the different microsampling devices. There is a decrease in signal compared to serum samples (Fig. 2D), which is likely due to a higher dilution factor after the sample extraction step. Nonetheless, we obtain a PPV of 93·01%, a NPV of 98·34% and a diagnostic accuracy of 96·98% considering a 25% prevalence (Table 1). The reproducibility between two measurements (N=143) on separate days again showed a good correlation with an R^2^ = 0·9655 (Fig. 3C). The percentage agreement with the Roche reference assay was 95·80% with a Kappa coefficient of 0·902 (95% CI: 0·825-0·978) (Table 1). This excellent agreement with the Roche serum analysis is promising as it takes into account all additional steps such as microsampling, dried shipping from one institution to another, manual extraction, freezing of the extract, and finally testing on NIA.

**Table 1:** Performance characteristics of microfluidic nano-immunoassay (NIA) index test. In all cases, the sensitivity and specificity values correspond to the use of the positivity threshold of the maximum likelihood ratio. For the comparison with the Roche reference standard, the same threshold is applied for category assignment. For the Euroimmun test, the positivity threshold recommended by the manufacturer (>1·1) is used. The number of agreements with the Roche reference assay are presented as well as the corresponding Kappa coefficient. Positive and negative predictive values (PPV and NPV) and accuracy are provided for a prevalence of 25%.

|  | Sensitivity | 95% CI | Specificity | 95% CI | PPV | NPV | Accuracy | Agreements | Kappa | 95% CI |
| --- | --- | --- | --- | --- | --- | --- | --- | --- | --- | --- |
| *Serum* |  |  |  |  |  |  |  |  |  |  |
| **Euroimmun** | **51.40%** | 44.12-58.61 | **100%** | 91.62-100.00 | **100%** | **86.06%** | **87.85%** | **60.63%** | **0.287** | 0.205-0.369 |
| **NIA** | **98.33%** | 95.22-99.55 | **97.62%** | 87.68-99.80 | **93.23%** | **99.43%** | **97.79%** | **98.20%** | **0.942** | 0.886-0.998 |
| *Microsampling* |  |  |  |  |  |  |  |  |  |  |
| **Mitra** | **95.05%** | 88.93-97.87 | **97.62%** | 87.68-99.88 | **93.01%** | **98.34%** | **96.98%** | **95.80%** | **0.902** | 0.825-0.978 |
| **Test strips** | **61.11%** | 51.69-69.77 | **100%** | 91.97-100.0 | **100%** | **88.52%** | **90.28%** | **72.55%** | **0.478** | 0.364-0.593 |
| **Hemaxis** | **83.16%** | 74.38-89.36 | **97.37%** | 86.51-99.87 | **91.33%** | **94.55%** | **93.82%** | **87.22%** | **0.720** | 0.600-0.840 |
| **Hemaxis DBS-A** | **91.49%** | 80.07-96.64 | **100%** | 81.57-100 | **100%** | **97.24%** | **97.87%** | **93.75%** | **0.851** | 0.711-0.991 |

**Figure 3:**
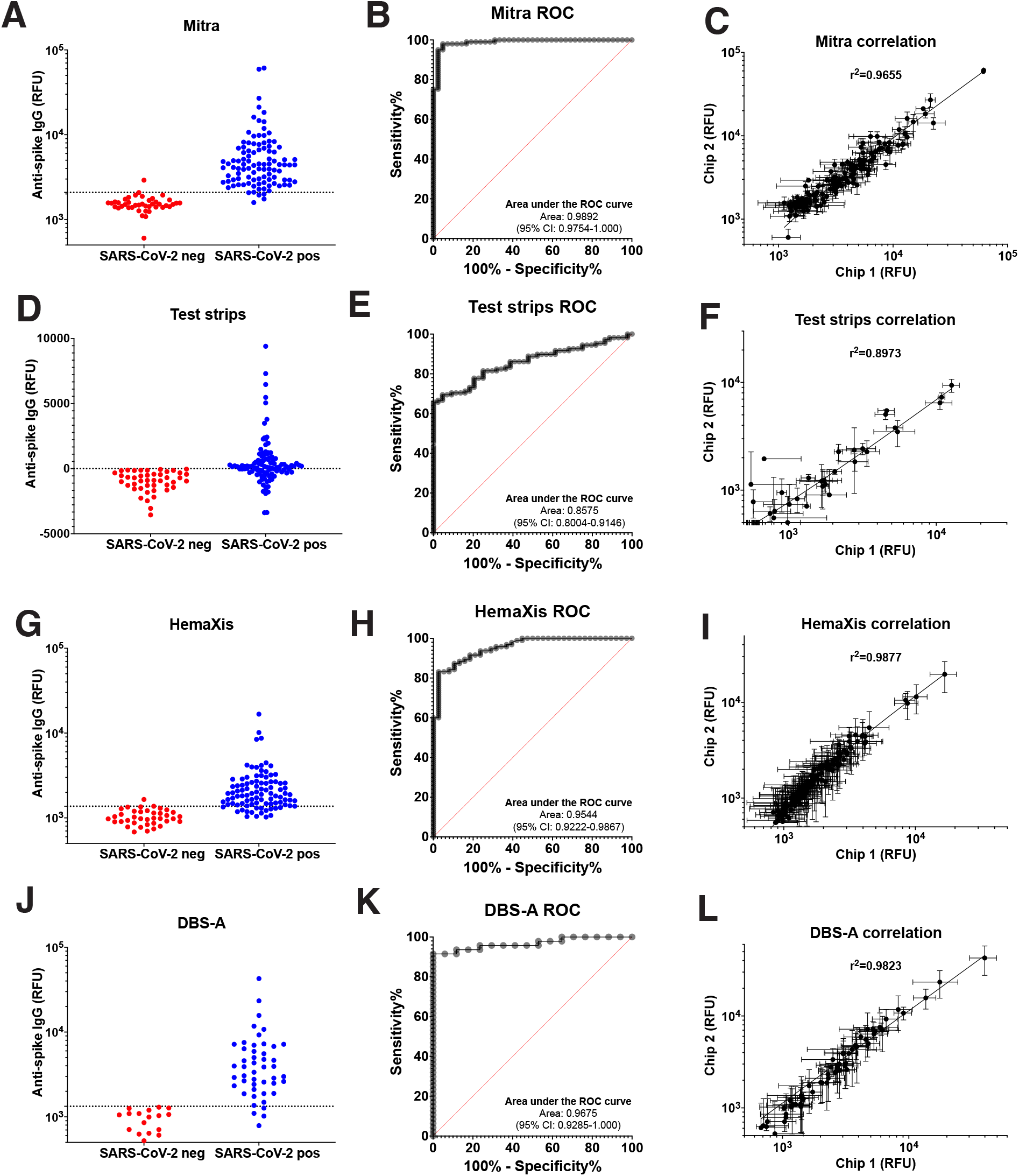
Performance of microfluidic nano-immmunoassay (NIA) test using dried blood microsamples. Signal intensities with positivity threshold (dotted line), ROC curve, and chip-to-chip correlation for the microfluidic nano-immunoassay testing of dried blood samples collected on Mitra **(A-C)**, (glucose) test strips **(D-F)**, or HemaXis microsampling devices **(G-I)**, and after automated extraction by flow-through desorption of dried blood spots (DBS-A) **(J-L)**.

### Glucose test strips

Using widely available and low-cost glucose test strips could potentially offer an alternative to currently available commercial devices, but the ultra-low volume of blood collected by glucose test strips is generally not sufficient for analysis with existing immunoassays. In our previous study, we showed as a proof-of-concept that it was technically feasible to measure small volume samples of venous blood sampled by the repurposed glucose test strips[22]. Here, combining capillary blood collection on glucose test strips with NIA showed modest discrimination between SARS-CoV-2 positive and SARS-CoV-2 negative samples (Fig. 3D). Indeed, the background-subtracted signal is noticeably lower compared to serum at 1:8 dilution or using the other microsampling devices, with a significant number of samples displaying higher background signal in the microfluidic chamber outside of the MITOMI assay region, resulting in negative background-substracted signals. The AUROC is 0·8575 (95% CI: 0·8004-0·9146) (Fig. 3E) and the positivity threshold selected as the lowest non-negative background substracted signal (>8·167 RFU) gives a clinical sensitivity of 61·11% (95% CI: 51·69-69·77) and specificity of 100% (95% CI: 91·97-100·0). This represents a PPV of 100%, a NPV of 88·52% and a diagnostic accuracy of 90·28% (Table 1). The higher dilution factor when extracting ultra-low sample volume of ∼0·6 *µ*L in 30 *µ*L buffer leads to a decrease in signal which negatively impacts sensitivity. While the decrease in signal would still allow identification of subjects with sufficiently high antibody levels, it might be more challenging in populations in which subject antibody levels have decreased due to a long interval since previous infection as we find here. The reproducibility between two separate measurements (N=152) showed a correlation with an R^2^ = 0·8973 (Fig. 3F), and the percentage agreement with the Roche assay was lower compared to Mitra at 72·55% and a Kappa coefficient of 0·478 (95% CI: 0·364-0·593) (Table 1).

### HemaXis device

Combining capillary blood collection on Hemaxis device with NIA showed good discrimination between SARS-CoV-2 positive and SARS-CoV-2 negative samples (Fig. 3G). The AUROC is 0·9544 (95% CI: 0·9222-0·9867) (Fig. 3H) and the threshold for positivity using the maximum likelihood (>1372 RFU) gives a clinical sensitivity of 83·16% (95% CI: 74·38-89·36) and specificity of 97·37% (95% CI: 86·51-99·87). We note a decrease in signal from samples collected on HemaXis compared to the samples collected on the Mitra device, even though the sampled (10 *µ*L) and extracted volumes (200 *µ*L) were identical. One possible explanation is that sample extraction is generally less efficient due to the different materials used, or that extraction is slower for HemaXis and would require more than the 3h extraction step used in this study. Indeed, we didn’t observe any difference in our previous study where samples were extracted overnight at 4^°^C [22]. At a prevalence of 25%, these results represent a PPV of 91·33%, a NPV of 94·55% and a diagnostic accuracy of 93·82% (Table 1). The reproducibility between two measurements (N=133) on different days showed a good correlation with an R^2^ = 0·9877 (Fig. 3C). The percentage agreement with the Roche assay was 87·22% with a Kappa coefficient of 0·720 (95% CI: 0·600-0·840) (Table 1).

### HemaXis device and automated flow-through desorption

As manual punching of dried blood spots from filter paper cards requires considerable operator time, we also tested the use of an automated dried blood spots (DBS-A) extraction method[25]. We observed that using flow-through desorption with a fully automated instrument (DBS-A, Gesrtel AG) improved the discrimination between SARS-CoV-2 positive and SARS-CoV-2 negative samples collected on filter paper cards with the Hemaxis device (Fig. 3J). The AUROC was 0·9675 (95% CI: 0·9285-1·000) (Fig. 3H) and the threshold for positivity using the maximum likelihood (>1333 RFU) returned a clinical sensitivity of 91·49% (95% CI: 80·07-96·64) and specificity of 100% (95% CI: 81·57-100). When comparing only with the subset of corresponding samples, manual extraction of HemaXis would present a clinical sensitivity of 87·50% (95% CI: 75·30-94·14) and specificity of 100% (95% CI: 81·57-100). While automated extraction still corresponds to a clinical sensitivity improvement and an increase in assay signal, it would be necessary to analyze a larger proportion of the samples for a more accurate comparison. Also, the confidence intervals for the sensitivity and specificity evaluation were relatively large due to the smaller number of samples analyzed. The higher clinical sensitivity and specificity using DBS-A extraction together with a prevalence of 25% corresponds to a PPV of 100%, a NPV of 97·24% and a diagnostic accuracy of 97·87%. Finally, the reproducibility between two separate measurements (N=65) showed a good correlation with an R^2^ = 0·9823 (Fig. 3C), and the percentage agreement with the Roche assay was 93·75% with a Kappa coefficient of 0·851 (95% CI: 0·711-0·991) (Table 1).

### Comparison between serum and dried blood samples

Previous findings have shown that anti-SARS-CoV-2 antibodies are reliably eluted from whole blood loaded on Mitra microsamplers and present equivalent signal compared to serum samples after testing on ELISA[16]. Also, NIA is capable of providing information on antibody concentration in serum using a single measurement at 1:8 dilution[22]. Here, we investigate the correspondence between signals obtained using microsampling and matched serum samples obtained by venipuncture on the same day.

We observed a modest correlation between serum samples and the dried blood microsample eluates tested on NIA with an R^2^=0·6779 for Mitra, R^2^=0·5568 for HemaXis, and R^2^=0·7217 for HemaXis after DBS-A extraction (Fig. 4 A, C, and D). On the other hand, using test strips for capillary microsampling leads to an absence of correlation with serum samples with an R^2^=0·1678(Fig. 4 B). Capillary blood sampling of a defined micro-volume using Mitra or HemaXis, together with complete microsample extraction helps in avoiding the hematocrit bias associated with partial dried blood spot processing[30]. However, variation in hematocrit among individuals[31] could still contribute to some variation in the antibody concentration in capillary microsamples and explain the modest correlation observed. We therefore assessed the correlation between the samples obtained on different microsampling devices. While the correlation remains low when comparing samples obtained using Mitra and glucose test strips with an R^2^=0·5414(Fig. 4 E), we see a good correlation between samples obtained using Mitra and HemaXis with an R^2^=0·7791 (Fig. 4 F), and an R^2^=0·8632 after DBS-A extraction (Fig. 4G). This observation suggests that hematocrit levels or factors influencing intra-individual differences between serum and capillary blood antibody levels could be in part responsible for the observed variation in signal on NIA.

**Figure 4:**
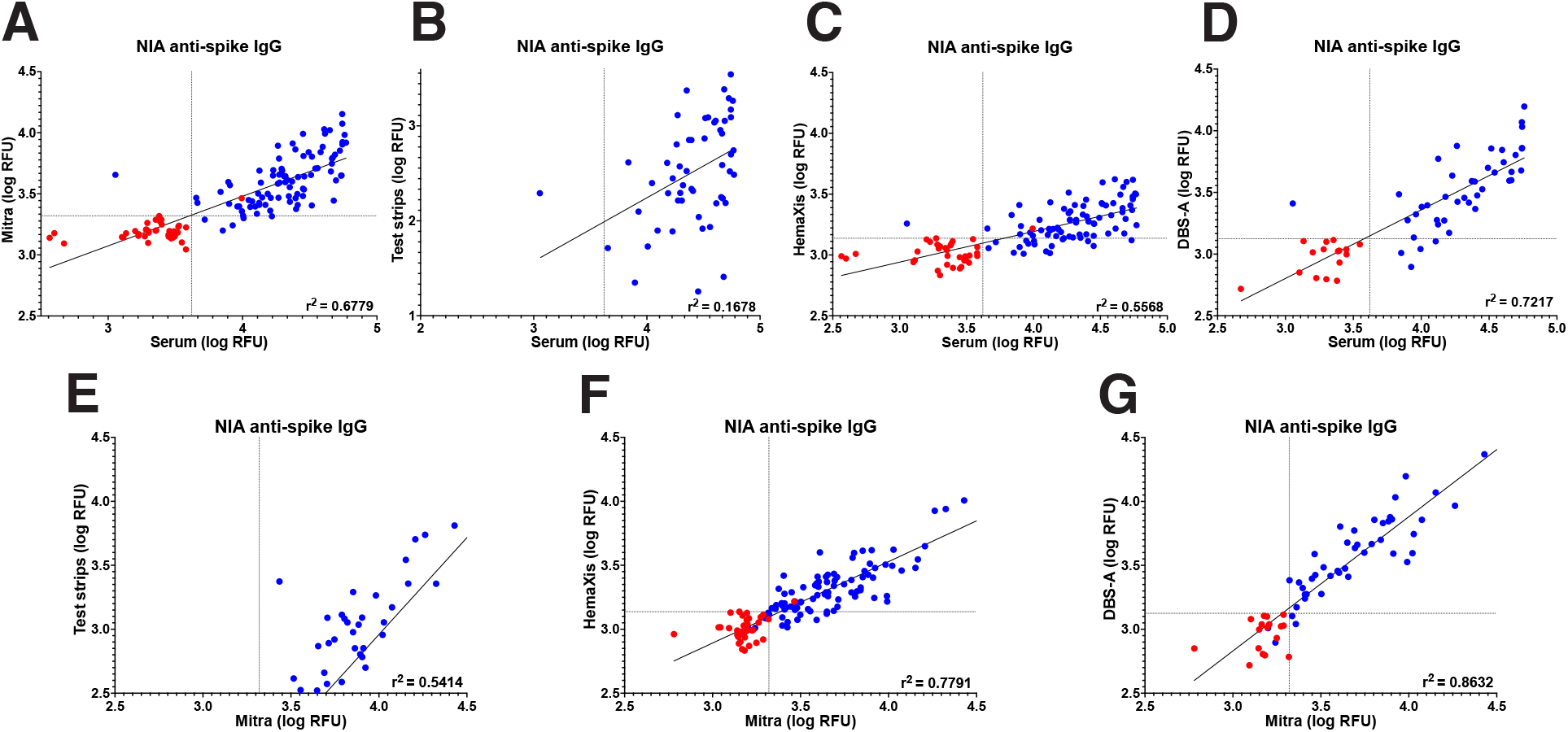
NIA serum and microsampling comparison. Comparison of serum at 1:8 dilution and samples obtained by microsampling tested on NIA. Samples from SARS-CoV-2 negative and SARS-CoV-2 positive participants are labeled red and blue, respectively. Comparison of serum samples with **A)** Mitra, **B)** (glucose) test strips, **C)** HemaXis, and **D)** HemaXis with automated extraction (DBS-A). Serum samples with signal above 59000 RFU were removed from the plot and linear regression. Comparison of Mitra with **E)** glucose test strips, **F)** HemaXis, and **G)** HemaXis with automated extraction (DBS-A). In panels **B)** and **F)**, samples with negative signals were excluded. The samples from SARS-CoV-2 negative participants were not included in the linear regression.

## Discussion

This study presents an extensive evaluation of micro-volume capillary blood sampling devices with analysis on a recently developed high-throughput microfluidic nano-immunoassay platform (NIA). Validation with serum samples confirmed the excellent performance of NIA. Of particular note is that samples in this study were obtained from participants with a long interval of 11 months between infection and blood sampling, which leads to decreases in antibody titers and therefore requires high-performance assay to return accurate results[26]. Moreover, the high performance of NIA on late time-point serum samples was obtained by measurements performed without any signal amplification, using only a few nanoliters of sample per assay, minute reagent consumption, and a throughput of 1024 assay points for each run on the microfluidic nano-immunoassay. While the clinical sensitivity of 98·33% in NIA was lower than the Elecsys anti-SARS-CoV-2 S total Ig reference assay, it was higher than the commercial anti-SARS-CoV-2 S1 IgG ELISA having only 51·40% clinical sensitivity at 11 months post infection when using the cutoff recommended by the manufacturer. This observation is confirming previous results showing a decrease in clinical sensitivity for serum samples obtained from individuals infected many months prior to sample collection, and also points out that selection of the appropriate test or reevaluation of the cutoff might be necessary depending on the intended use of the assay, such as when performing serological surveys[26].

We also note that the antigen and format used in the different assays might lead to some differences in the seropositivity evaluation, as the Elecsys anti-SARS-CoV-2 S total Ig assay uses RBD antigen in a sandwich format detecting all antibody classes, while the anti-SARS-CoV-2 S1 IgG ELISA uses the S1 part of the spike protein immobilized on a plate and detects IgG antibodies only. On the other hand, NIA uses a full-length trimerized spike pulled down on the assay surface by an anti-His-tag antibody and detects IgG antibodies.

Following validation using serum samples, evaluation of the combined use of NIA with samples obtained by capillary blood microsampling demonstrated its suitability for serological surveys. First, testing of capillary blood obtained by fingerprick using a Mitra microsampling device and analyzed with NIA offers performance characteristics comparable to tests performed in the laboratory using serum obtained by venipuncture, with a clinical sensitivity of 95·05% in participants 11 months post infection. The performance is better than a lateral flow immunoassay (LFIA) evaluated[32] and selected for a large-scale seroprevalence study[33], or other more recently developed LFIAs [34], which shows the utility of Mitra microsampling combined with NIA in conducting decentralized serological surveys.

Using glucose test strips, the clinical sensitivity is lower than using purpose built and professionally designed microsampling devices. However, with their low cost and wide availability, repurposed glucose test strips could play a role when conducting large serosurveillance studies, provided improvements can be made in the sampling or extraction parameters to allow sufficient detection of antibody signal in such challenging late time-point samples. On the other hand, the use of glucose test strips for early time-point samples is likely already of sufficient quality based on our previous results[22], although this remains to be assessed in a suitable cohort.

Collection of capillary blood on HemaXis and testing on NIA affords relatively good sensitivity and specificity when analyzed on NIA. Although the HemaXis device showed lower sampling success rates compared to Mitra or glucose test strips, the success in sampling is still relatively high at 86,2%. An advantage of using HemaXis compared to Mitra device is the wide use of filter paper for sample collection and the associated methods for processing the dried blood samples, such as automated punchers which are commonly used in neonatal screening programs[35]. To benefit from HemaXis volumetric sampling and to avoid hematocrit bias, it is however preferable to extract the entire dried blood spot. While manual punching and extraction offered lower sensitivity when compared to results obtained with Mitra, flow-through desorption of the entire dried blood spot using a fully automated dried blood spot extraction (DBS-A) improved the clinical performance. With barcode recognition capability and robotic processing, an efficient workflow can be envisioned in combination with the high-throughput of NIA.

We recognize different limitations in our study and acknowledge the following explicitly. We relied for the SARS-CoV-2 positive group on participants already involved in a longitudinal evaluation of anti-SARS-CoV-2 antibody levels and all participants had a positive result on serology at the baseline visit. While the serological status at the baseline did not constitute a selection criterium for the SARS-CoV-2 positive group, this can lead to an overestimation of the sensitivity as the SARS-CoV-2 positive group does not include potential patients with previous infection who did not develop an antibody response[26]. The participants were all presenting mild to moderate symptoms and no asymptomatic patients, nor patients with severe or critical illness requiring further care were included. The participants in the SARS-CoV-2 negative group formed a convenience sample which might not be fully representative of the general population, and 5 participants from the SARS-CoV-2 negative group had to be excluded from the study due to positive signal on the Roche reference assay. The specificity evaluation did not address some common interference in immunoassays such as for patients with autoimmunity or chronic inflammation, and could be investigated further in a study including such patients. Moreover, we did not test cross-reactivity to other coronaviruses or other respiratory viruses which is another potential cause of false positives. Finally, the number of participants was not high, and resulted in relatively broad confidence intervals for the specificity evaluation (Fig. 5), but also for the sensitivity evaluation when the observed sensitivity was lower than expected from previous results[22].

**Figure 5:**
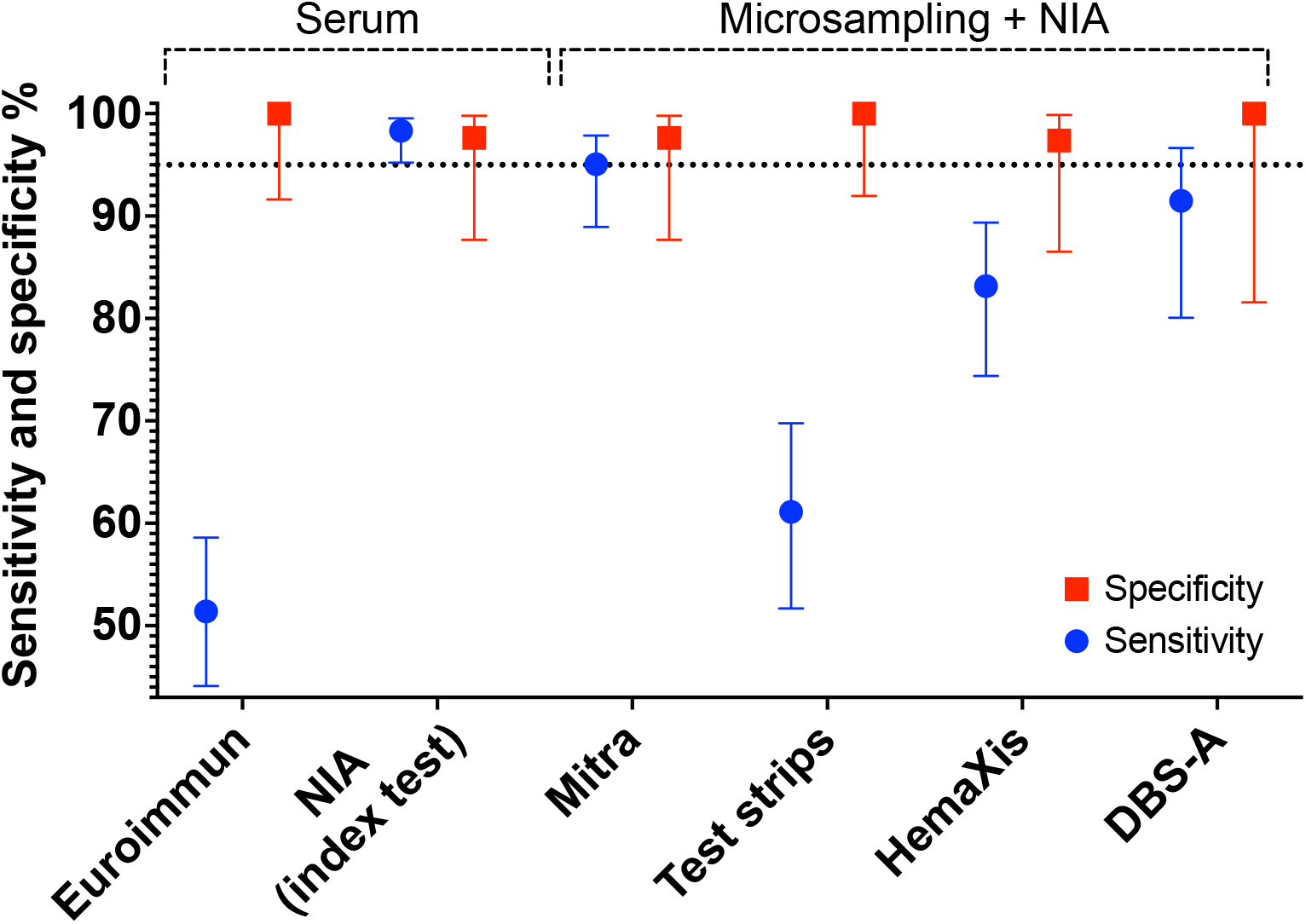
Sensitivity and specificity comparison. Sensitivity and specificity with 95% confidence interval are shown. Euroimmun ELISA and NIA index test were performed on serum obtained by venipuncture. Mitra, (glucose) test strips, and HemaXis correspond to the device used for capillary blood collection after fingerprick, and DBS-A to automated extraction of samples obtained on HemaXis. Capillary blood samples were dried, shipped, and extracted before testing on the NIA assay. A dotted line at 95% sensitivity or specificity is shown for visual aid.

In conclusion, we evaluated in this study micro-volume capillary dried blood sampling methods combined with a high-throughput microfluidic nano-immunoassay (NIA)[22] and showed its potential for use in conducting serological surveys for COVID-19. In a cohort with SARS-CoV-2 positive participants 11 months post infection, the high clinical sensitivity and specificity of NIA was demonstrated when testing serum samples as well as dried blood microsamples obtained by fingerprick, which should facilitate its use in large-scale studies using home-based sampling. The simplified sample collection will also be useful when testing vulnerable populations where blood sampling by venipuncture is difficult to implement, such as in the elderly[36] or pediatric populations[37, 38]. With its high-throughput, reduced reagents and low sample consumption, the platform might benefit vaccine research in establishing remote monitoring of participants, which could facilitate logistics and reduce the costs associated with such clinical trials. Finally, the open design of NIA allows for the rapid modification of the assay allowing to perform serological surveys and gather information on present and future SARS-CoV-2 variants, other circulating pathogens of public health importance, as well as emerging infectious diseases.

## Data Availability

All data included in the article and corresponding data will be made available as supplementary files with publication.

## Author contributions

SJM, BM, and IE developed the project. MB and PS supervised sample collection using microsampling. GM, FA, OP performed experiments. AGL performed experiments, provided resources. GM, IE, BM, SJM, designed experiments, analyzed data, and wrote the manuscript with input from all authors.

## Declaration of interests

The automated DBS extraction was performed using an instrument loaned free of charge by Gerstel AG. Gerstel AG did not take part in the decision to publish the study and did not have editorial control of the results presented.

## Acknowledgments

We would like to thank the participants of the study and the nursing staff for their help in the collection of blood samples. We thank Florence Pojer, Kelvin Lau, and David Hacker from the EPFL Protein Production Facility for providing us with the purified SARS-CoV-2 spike protein. We thank Pauline Vetter, Carole Salomon, and the Clinical Research Center of the Geneva University Hospital. We thank Laurent Kaiser, Isabelle Arm-Vernez, and the Laboratory of Virology of the Geneva University Hospital for the technical assistance. This work was supported by the SNF NRP 78 Covid-19 grant 198412 (SJM, IE, and BM), and a grant from the Private Foundation of the Geneva University Hospital (IE).

## Notes

### Author Declarations

Ethical approval for this study was given by the Ethics Committee Geneva (Commission Cantonale d'Ethique de la Recherche sur l'etre humain (CCER) project numbers 2020-00516 and 2020-02323) and registered (NCT04329546) prior to initiation.

